# Continued underutilization with pronounced geographic variation in clozapine use

**DOI:** 10.1101/2024.04.08.24305459

**Authors:** Luke R. Cavanah, Maria Y. Tian, Jessica L. Goldhirsh, Leighton Y. Huey, Brian J. Piper

## Abstract

**Introduction:** Schizophrenia-spectrum disorders are debilitating and contribute to a substantial economic burden. Clinicians have historically underutilized clozapine, an atypical antipsychotic traditionally reserved for use in treatment-resistant schizophrenia, due to the medication’s adverse effect profile and associated management requirements, concerns of poor treatment adherence, and poor training/exposure to the use. In addition to alleviating schizophrenia symptoms when multiple other medications have failed, clozapine has other unique benefits that compel its use such as its use being associated with reduced suicide ideation and action, aggression, substance use, and all-cause mortality.

**Methods:** This study aimed to characterize clozapine utilization by US Medicare patients from 2015-20. Additionally, we identified the states that prescribed significantly different amounts than the national average.

**Results:** We observed a steady decrease in clozapine use adjusted for population (−18.0%) and spending (−24.9%) over time. For all years, there was significant geographic heterogeneity (average: nine-fold) in population-corrected clozapine use. Massachusetts (2015-20: 95.4, 82.7, 76.8, 72.2, 71.2, 63.7 prescriptions per thousand enrollees) and South Dakota (2015-20: 78.0, 77.4, 78.4, 75.6, 72.0, 71.6) were the only states that prescribed significantly more than average, and none prescribed significantly less.

**Discussion:** Clozapine use by US Medicare patients is low, decreasing, and concerning for underutilization—patterns likewise seen for the US Medicaid recipients. Further study of the reasons for the state variation is needed. Education interventions, training reform, and devices that ease required routine blood monitoring are all practical solutions to optimize clozapine use.

## Introduction

Schizophrenia-spectrum disorders include chronic psychiatric disorders characterized by episodes of psychosis, disorganized thought, and a decline in functioning (American Psychiatric Association, 2022). As of 2016 in the US, age-standardized point prevalence and burden of schizophrenia were estimated to be 0.30% and 191.5 years lost to disability per hundred thousand, respectively (Charlson et al., 2018). About one-third of cases of schizophrenia are deemed treatment-resistant (TRS), meaning two or more antipsychotics (APs) of adequate dose, trial length, and adherence failed to resolve the symptoms (Meltzer, 1997). Even with the low prevalence, schizophrenia results in a massive and multifaceted burden: reduced quality of life, cognitive and social functioning, and lifespan; increased depression, substance use disorder, medical comorbidities, need for a caregiver, and increased likelihood of experiencing unemployment and homelessness (Millier et al., 2014). Further, schizophrenia negatively impacts the psychosocial and financial aspects of the families and caregivers (Millier et al., 2014). Economically, the magnitude of the burden is similarly large and diverse, costing the US $155.7 billion, 24% of which is due to direct healthcare costs, 6% to direct non-healthcare costs, and 76% to indirect costs (Cloutier et al., 2016). TRS is estimated to cost 3-11-fold more than schizophrenia which is treatment-responsive (Kennedy et al., 2014).

Clozapine is a second-generation (atypical) AP that is indicated for the treatment of TRS and the related treatment-resistant schizoaffective disorder as well as suicidality in schizophrenia-spectrum disorders (American Psychiatric Association, 2020; Barnes & Schizophrenia Consensus Group of British Association for Psychopharmacology, 2011; National Institute for Health and Care Excellence, 2015; Royal Australian and New Zealand College of Psychiatrists Clinical Practice Guidelines Team for the Treatment of Schizophrenia and Related Disorders, 2005). Clozapine’s mechanisms of action include serotonergic blockade, weak dopamine antagonism, and potent muscarinic, histamine, and alpha-1 adrenergic antagonism (Stahl, 2021). Not only is clozapine an effective medication when first- and second-line treatments have failed (J. Kane et al., 1988), it is the only indicated and evidence-based treatment for TRS (American Psychiatric Association, 2020; Barnes & Schizophrenia Consensus Group of British Association for Psychopharmacology, 2011; J. M. Kane et al., 2019; National Institute for Health and Care Excellence, 2015; Royal Australian and New Zealand College of Psychiatrists Clinical Practice Guidelines Team for the Treatment of Schizophrenia and Related Disorders, 2005). Even more, a recent meta-analysis and systematic review indicates that clozapine may be more effective than other AP even when used as first- or second-line treatment for schizophrenia (Okhuijsen-Pfeifer et al., 2018), which is an important finding since there is some speculation that TRS is a neurobiologically distinct disorder from treatment-amenable schizophrenia (Demjaha et al., 2014; Potkin et al., 2020).

In addition to being the only established effective treatment for TRS (American Psychiatric Association, 2020; Barnes & Schizophrenia Consensus Group of British Association for Psychopharmacology, 2011; J. M. Kane et al., 2019; National Institute for Health and Care Excellence, 2015; Royal Australian and New Zealand College of Psychiatrists Clinical Practice Guidelines Team for the Treatment of Schizophrenia and Related Disorders, 2005) and having superior efficacy compared to other medicines in its class (Okhuijsen-Pfeifer et al., 2018), clozapine has other benefits that warrant attention and possibly other uses. Clozapine has a strongly supported anti-suicide effect in schizophrenia/schizoaffective disorder patients, and there even is some evidence that is has similar effect in severe and treatment-resistant suicidality and non-suicidal self-injury (NSSI) in patients with bipolar disorder or borderline personality disorder (Masdrakis & Baldwin, 2023). The anti-suicide benefit of clozapine is important because patients with schizophrenia, TRS, bipolar disorder, and borderline personality disorder all have significantly elevated suicide risk (Dome et al., 2019; Hor & Taylor, 2010; Pompili et al., 2005). The possible NSSI benefit is important because it is a strong predictor of suicide (Karasouli et al., 2015), about one-fifth of people will engage in NSSI at some point in their life (Muehlenkamp et al., 2012; Swannell et al., 2014), it increases the risk of suicide (Olfson et al., 2018), and is refractory to numerous psychological and pharmacological therapies (Brausch & Girresch, 2012). Another unique but less supported benefit of clozapine is reduced aggression in patients with schizophrenia-spectrum disorders (Frogley et al., 2012).

Despite all these advantages, clozapine utilization remains limited by concerns of serious adverse effects, mandatory blood tests, perceived likelihood of patient noncompliance, clinician challenges in recognizing appropriate patients, and insufficient training and exposure to clozapine therapy (Farooq et al., 2019; Meltzer, 2012). Like other atypical antipsychotics, clozapine has the risk of metabolic syndrome, constipation (14-25%), orthostatic hypotension, sedation (≤46%), sexual dysfunction, and extrapyramidal symptoms (EPS) (Novartis Pharmaceuticals Corporation, 1989). Of note, clozapine has a higher risk of metabolic syndrome (Pillinger et al., 2020) and the lowest risk of EPS (Divac et al., 2014; Strejilevich et al., 2005) relative to other pharmaceuticals in its class. The most concerning adverse effects of clozapine, however, are leukopenia (≤3%) and agranulocytosis (1-2%) (Mijovic & MacCabe, 2020; Novartis Pharmaceuticals Corporation, 1989), which can lead to life-threatening infections. Cardiovascular adverse effects include dose-independent myocarditis and cardiomyopathy (Novartis Pharmaceuticals Corporation, 1989). Neurologic adverse effects include a dose-dependent risk of seizures (Novartis Pharmaceuticals Corporation, 1989). The hematologic risk of clozapine use requires practitioners to place patients taking clozapine on a national registry and to frequently monitoring of white blood cell and absolute neutrophil counts (American Psychiatric Association, 2020). In spite of clinician concerns of clozapine toxicity, especially the potentially fatal effect of agranulocytosis, clozapine use is associated with decreased all-cause mortality risk (Cho et al., 2019).

Clozapine use has a historic pattern in countries throughout the world of being low and incongruent with guidelines (Bachmann et al., 2017; Benito et al., 2023; Ismail et al., 2019; Latimer et al., 2013; Nielsen et al., 2012; Stroup et al., 2014; Warnez & Alessi-Severini, 2014; Whiskey et al., 2021). Contrary to the patterns of increased prevalence of clozapine in most populations examined by a recent study of international trends in clozapine use, there was a significant decline in clozapine utilization in publicly insured U.S.—the only other decline seen was by Columbia (Bachmann et al., 2017). Of note, however, the publicly insured US population was one of few groups in the study which prescribed the amount of clozapine consistent with crude estimates of individuals with TRS, granted this crude estimation neglects clozapine use outside of TRS (Bachmann et al., 2017). If clinicians are not using clozapine when they encounter TRS, it begs the question, what are they doing? Strategies commonly applied in such situations include increased trial duration and/or dosage, trial of another non-clozapine AP, and augmentation with non-clozapine APs and/or mood stabilizers, all of which are practices with scant supporting evidence and may lead to unnecessary risk (Agid et al., 2007, 2011, 2013; Correll et al., 2017; Galling et al., 2017; Howes et al., 2012; Patel et al., 2014; Paton et al., 2008; Üçok et al., 2015; Walkup et al., 2000). Delaying clozapine treatment might not only mean prolonged symptomatology due to time spent on treatments with little efficacy, but it also can reduce the effectiveness of clozapine when it is finally tried (Yoshimura et al., 2017).

While clozapine use is nearly uniformly underutilized, its rate of use appears to vary substantially from region to region within a country (Benito et al., 2023; Ismail et al., 2019; Latimer et al., 2013; Nielsen et al., 2012; Stroup et al., 2014; Whiskey et al., 2021) and from country to country (Bachmann et al., 2017). More specifically, there has been considerable geographic variation of clozapine use in the US Medicaid system, with the most recent study showing a 13.0-fold state-level difference in 2019, with South Dakota, Missouri, and North Dakota prescribing significantly more than the national average, and not states prescribing significantly less (Benito et al., 2023). Though high state-level variation within the US Medicaid system has been observed in three recent epidemiologic investigations of clozapine and many more by other psychotropics, such as esketamine, methadone, and buprenorphine (Aguilar et al., 2023; Dana et al., 2024; Kennalley et al., 2023), few reasons have been set forth to understand the reasoning behind this. Like Medicaid, pronounced state variation in U.S.

Medicare system has been noted in the prescription patterns of psychotropics, such as Z-drugs, and buprenorphine (Anderson et al., 2023; Hsu et al., 2023), but no recent investigations have examined this for clozapine.

In summary, use of clozapine, the only indicated and empirically supported treatment for TRS, has been low and incongruent with recommendations due to the medicine’s perceived toxicity, required blood monitoring, and lack of training in recognizing candidates for clozapine and managing patients treated with clozapine. Considering the superior efficacy, reduced risk of suicidality and tardive dyskinesia associated with other APs, recognized effectiveness in TRS, and established history of suboptimal and geographically variable utilization of this life-saving medication, there is a need to examine the pharmacoepidemiologic patterns in clozapine use.

This study aimed to analyze the Medicare data from 2015-2020 to characterize the national and regional variation of clozapine use.

## Methods

### Procedures

Prescription rates and spending for clozapine were obtained for 2015-2020 for Medicare Part D patients. Prescriptions were defined as the number of Medicare Part D claims, and spending data was defined as drug cost paid for all associated claims. State-level data were extracted from Medicare Specialty Utilization and Payment Data (Centers for Medicare & Medicaid Services, n.d.). The study was approved as exempt by the Geisinger IRB.

### Data Analysis

Patterns of prescriptions, per thousand enrollees, and spending for clozapine were compared for generics, brands, and their total. States’ annual population-corrected prescriptions and the average cost of prescription were then compared to the mean for that respective year, and values outside a 95% confidence interval were considered significantly different. These methods have been used in numerous prior pharmacoepidemiologic reports to investigate geographic variation (Aguilar et al., 2023; Anderson et al., 2023; Benito et al., 2023; Dana et al., 2024; Hsu et al., 2023; Kennalley et al., 2023). Population-corrected prescription distribution in Medicare was compared to Medicaid in 50 US states for 2019 <u>(Benito et al., 2023)</u> by a correlational analysis). Data were analyzed and visualized using Excel, GraphPad Prism, and Heatmapper (Babicki et al., 2016).

## Results

### National prescribing

Figure 1 illustrates that both spending and utilization (corrected for population) gradually declined from 2015-2020. There were 32.2 prescriptions per thousand Medicare Part D enrollees in 2015, for which 164.3 million dollars was spent. In 2020, prescriptions per thousand enrollees were reduced by 18.0% to 26.4, while spending was reduced by 24.9% to 123.4.

**Figure 1.**
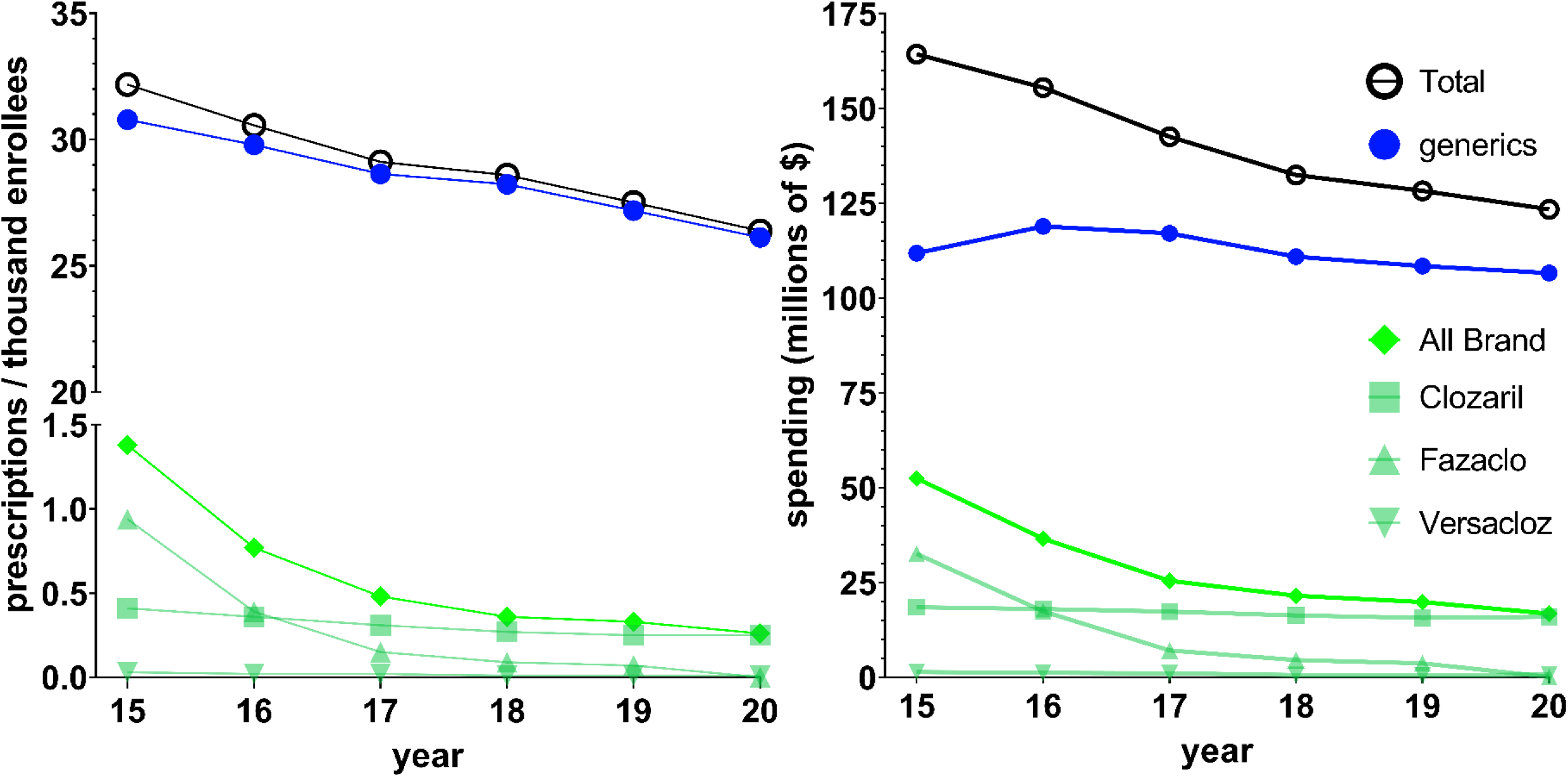
National Medicare prescription rates per thousand Part D enrollees (left) and Medicare spending (right) for clozapine gradually decreased for 2015-2020.

There was a decrease of 4.1% in prescriptions per thousand enrollees pre-COVID (2019) to post-COVID (2020). Moreover, Figure 1 also illustrates that the observed gradual decrease in utilization and spending applies to generics and all brand formulations. Generic clozapine accounted for the vast majority (95.7%) of prescriptions in 2015 and this increased to 99.0% in 2020. Spending on the brand was disproportionate to its use for 2015-2020. Brand name formulations comprised 31.9% of spending in 2015 and decreased to 13.6% by 2020.

### State prescribing

Figure 2 and Supplemental Figures 1-5 show that, throughout the decline in spending and population-corrected use of clozapine, there existed marked variability in 2015 (11.1-fold), 2016 (10.2-fold), 2017 (8.6-fold), 2018 (8.6-fold), 2019 (8.5-fold), and 2020 (8.6-fold) from one state to another. Massachusetts was the largest prescriber in 2015 (95.4 prescriptions per thousand enrollees) and 2016 (82.7), and South Dakota was the largest prescriber in 2017 (78.4), 2018 (75.6), 2019 (72.0), and 2020 (71.6). Both Massachusetts and South Dakota prescribed significantly more than average for all years examined. Additionally, New Hampshire prescribed significantly more than average in 2015 (76.5). No state prescribed significantly less than average although some states were below the 1.5 SD cutoff.

**Figure 2.**
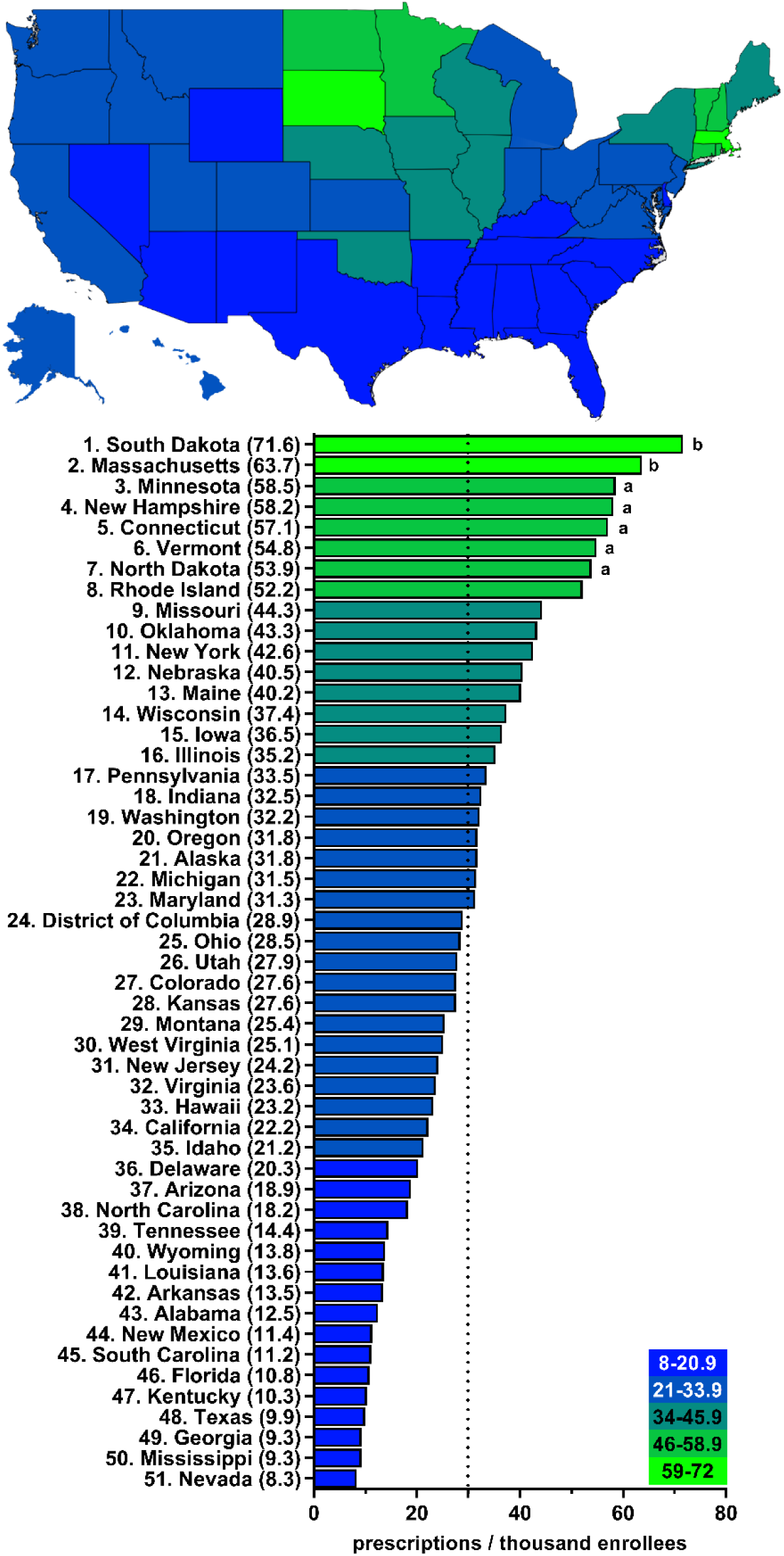
Large state-wide variation (8.6-fold) in number of clozapine prescriptions per thousand Medicare Part D enrollees in 2020 ^a^ indicates >1.50 SD (16.0) from the mean (29.9), which is denoted by the dotted line. ^b^ indicates >1.96 SD from the mean.

In addition to the pronounced state-state heterogeneity in the population-corrected number of prescriptions, Figure 3 shows there also existed significant state-wide variation (2.1-fold) in the average cost of a prescription in 2020. Supplemental Figures 8-14 reveal the presence of high state-wide variation was also present in 2015 (3.1-fold), 2016 (2.4-fold), 2017 (2.1-fold), 2018 (2.4-fold), and 2019 (2.2-fold). The average prescription of clozapine cost, of all states, was the most in Texas for 2015 ($202.00), 2016 ($187.02), 2017 ($165.55), and 2019 ($144.51), and cost significantly more than average from 2015-2019. In 2018, clozapine cost the most in Wyoming ($160.19), which was also the only year when it was significantly higher than the state average. In addition to Texas, the cost was often significantly higher for California: 2015 ($184.39), 2016 ($175.32), 2017 ($149.51), and 2020 ($140.72). Occasionally, significantly higher prices were seen in Nevada (2016: $177.18, 2019: $142.75), Delaware (2019: $144.08), and New Jersey (2020: $127.12). On the other end of the spectrum, the average cost was significantly lower in Alaska in 2015 ($65.63) and Hawaii in 2018 ($66.23).

**Figure 3.**
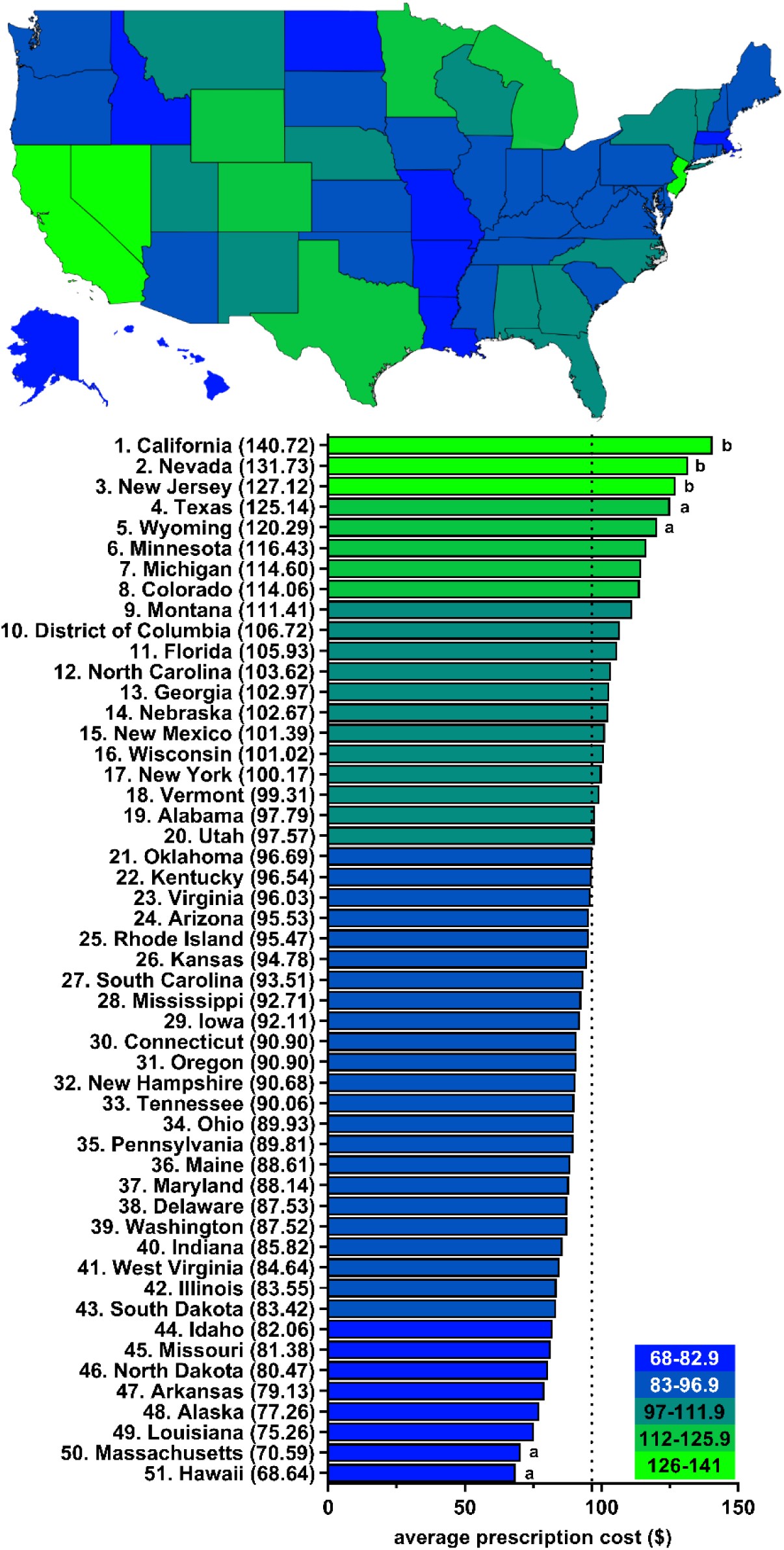
Pronounced state-wide variation (2.1-fold) in the average cost of a clozapine prescription for Medicare Part D enrollees in 2020. ^a^ indicates >1.50 SD ($15.08) from the mean ($96.48), which is denoted by the dotted line. ^b^ indicates >1.96 SD from the mean.

The prescription rates between Medicare and Medicaid correlated strongly (r(50) = 0.73, p <0.0001, Figure 4). In both Medicaid and Medicare programs in 2019, South Dakota prescribed the most, exceeding the average of Medicaid state prescriptions by 59.3% and Medicare state prescriptions by 57.1% (Figure 4). In Medicaid, Arkansas had the lowest prescription of 14.8 (Figure 4). In Medicare, Mississippi had the lowest prescription of 8.4 (Figure 4).

**Figure 4.**
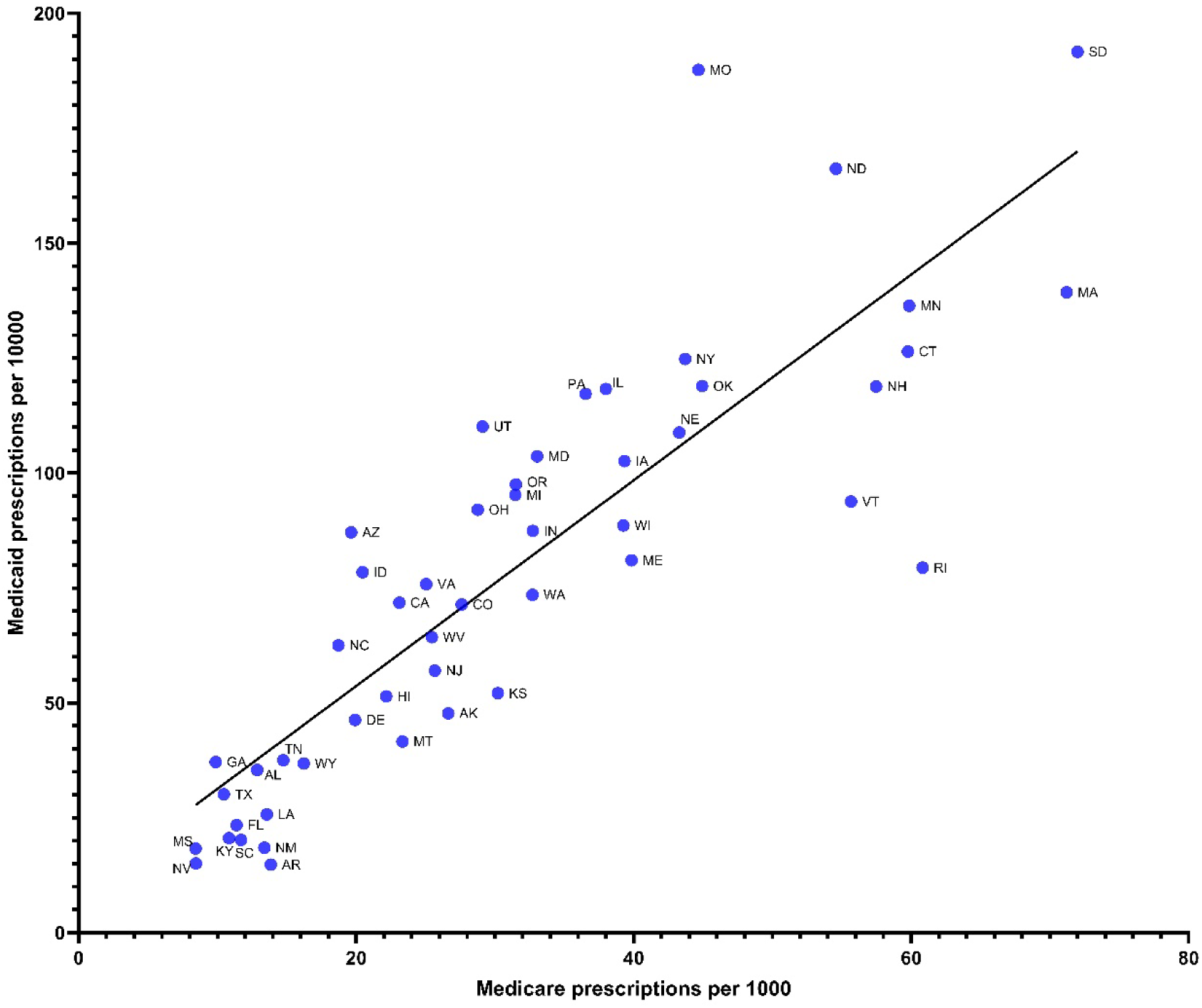
Strong correlation (R^2^ = 0.734 p < .0001) between Medicare prescriptions per 1,000 persons and Medicaid prescriptions per 10,000 persons among the fifty US states in 2019.

## Discussion

### National prescribing

There were two key findings from this novel study. First, as hypothesized, clozapine use in Medicare patients, like that of Medicaid patients (Benito et al., 2023), was low and decreasing (−18.0%) from 2015-2020. Further, the low clozapine use is suggestive of clozapine underutilization, a finding noted in the previous studies including the US and other nations (Bachmann et al., 2017; Benito et al., 2023; Ismail et al., 2019; Latimer et al., 2013; Nielsen et al., 2012; Stroup et al., 2014; Warnez & Alessi-Severini, 2014; Whiskey et al., 2021). The literature previously has attributed clozapine underutilization to clinician’s concerns of toxicity and patient non-adherence, requirement for patients to be placed on national registry and undergo routine blood monitoring, under-recognition of appropriate clozapine candidates, and poor training and exposure to clozapine use (Farooq et al., 2019; Meltzer, 2012), which is likely the underlying explanation for these present findings as well. Other reasons clinicians may be less likely to use clozapine is its relatively higher anticholinergic properties (Stahl, 2021) and higher risk of metabolic syndrome (Pillinger et al., 2020).

Since clozapine underutilization has been a historically consistent phenomenon that has substantial consequences for patients with psychotic disorders, their families and caregivers, and the economy at large (Cloutier et al., 2016; Kennedy et al., 2014; Millier et al., 2014), many studies have proposed solutions to enhance the use of clozapine to better match the recommendations of clinical guidelines (Bogers et al., 2015; Cohen & Farooq, 2020; Taylor et al., 2021). First and perhaps the most obvious solution to this problem is through education.

Education with emphasis on clozapine’s comparative efficacy, anti-suicide effects, anti-aggression benefits, and lower risk of EPS are all reasonable adjustments to psychiatric training. Introduction in psychiatric residency training of a mandatory training program with certification for clozapine treatment initiation and maintenance or exposure to clozapine clinics could yield important improvements in clozapine use (Cohen & Farooq, 2020; Olivia Freudenreich et al., 2013). A second possible solution for remedying clozapine under-prescription is the development of clinical tools that ease the burden of the associated medical management, especially the routine blood work. Solutions that have been tried and seem promising are tools that allow the measurement of clozapine plasma concentrations and white blood cell count through finger-stick capillary sample, like how patients with diabetes can monitor their blood sugar (Bogers et al., 2015; Taylor et al., 2021).

Although these findings are concerning for clozapine underutilization by Medicare Part D enrollees, the available Medicare data does not include the indication for why clozapine was used. Future studies that differentiate epidemiologic patterns by indication of clozapine use would overcome this limitation. Additional future directions that could augment this study’s findings pertaining to chronological variation include (1) estimating how the humanistic and economic burden of TRS would be reduced if clozapine use aligned with clinical guidelines and (2) to characterize the chronological variation of other treatment strategies for TRS.

### State prescribing

The second key finding was that, as hypothesized, clozapine use in Medicare patients, like that of Medicaid patients (Benito et al., 2023), exhibited substantial and significant geographic variability (average of nine-fold difference) from 2015-2020. The use of clozapine also correlated strongly among the fifty US states between Medicare and Medicaid (Figure 4). (Bachmann et al., 2017; Benito et al., 2023; Ismail et al., 2019; Latimer et al., 2013; Nielsen et al., 2012; Stroup et al., 2014; Whiskey et al., 2021)￼ their origins are complex and not well-understood. Patient characteristics associated with clozapine initiation include:(Stroup et al., 2014)￼.

Interestingly, although there was a similar magnitude of geographic variation within the US Medicaid population (Benito et al., 2023), the states that prescribed significantly different amounts than average were partially distinct. For Medicare, South Dakota and Massachusetts were the states that prescribed significantly more than the mean, and for Medicaid, South Dakota, Missouri, and North Dakota prescribed significantly more than average (Benito et al., 2023). The consistency of South Dakota prescribing significantly more than average for both insurance systems could mean that identification of the reasons for this finding could provide insight on how other states can optimize the use of clozapine. Rhode Island and Vermont were states that prescribed significantly more than average in Medicare but much less in Medicaid. Interestingly, an increase in a biased distribution of clozapine through Medicaid or Medicare was observed in states that had increasing prescription uses (Figure 4). There does not seem to be an overall increase of clozapine use through Medicaid or Medicare but future directions may be beneficial to explore factors that lead a state to prefer distributing drugs through one medical insurance program over another (Figure 4). To develop more robust explanations of the geographical variation observed by clozapine and other psychotropic medications, it would be helpful to conduct exploratory analyses with potential causes, such as percent white population (given concern for benign ethnic neutropenia), existence of clozapine clinics, number of psychiatrists and other psychiatric prescribers. Another approach to increase understanding of the geographic variation would be to investigate how the clinician’s attitudes toward clozapine (e.g., perceived toxicity, concerns of patient non-adherence) vary from one state to the next and conduct correlational analyses with state use. Qualitative studies of patients’ perspective might also be valuable. As also noted above, since the indication of clozapine is not included in the present study’s dataset, it would be valuable to stratify number of state population-corrected prescriptions by clozapine indication. Epidemiologic investigation of the use of other effective and underutilized psychiatric treatments and comparison of patterns to the present study’s findings may also reveal insights for possible reasons for underuse.

## Conclusion

In summary, rates of clozapine prescriptions to Medicare patients have decreased over the past six years, despite prior evidence of it having already being underutilized. Throughout this chronological decrease in prescriptions of clozapine, there was clear evidence of large (nine-fold) state-level variation. Future studies should explore possible associations underlying the decreasing and variable use of clozapine among Medicare patients.

## Data Availability

All data produced in the present work are contained in the manuscript

## Acknowledgments

BJP was supported by HRSA (D34HP31025). NIEHS (T32-ES007060-31A1) provided the software for this research.

---

**Supplemental Figure 1.**
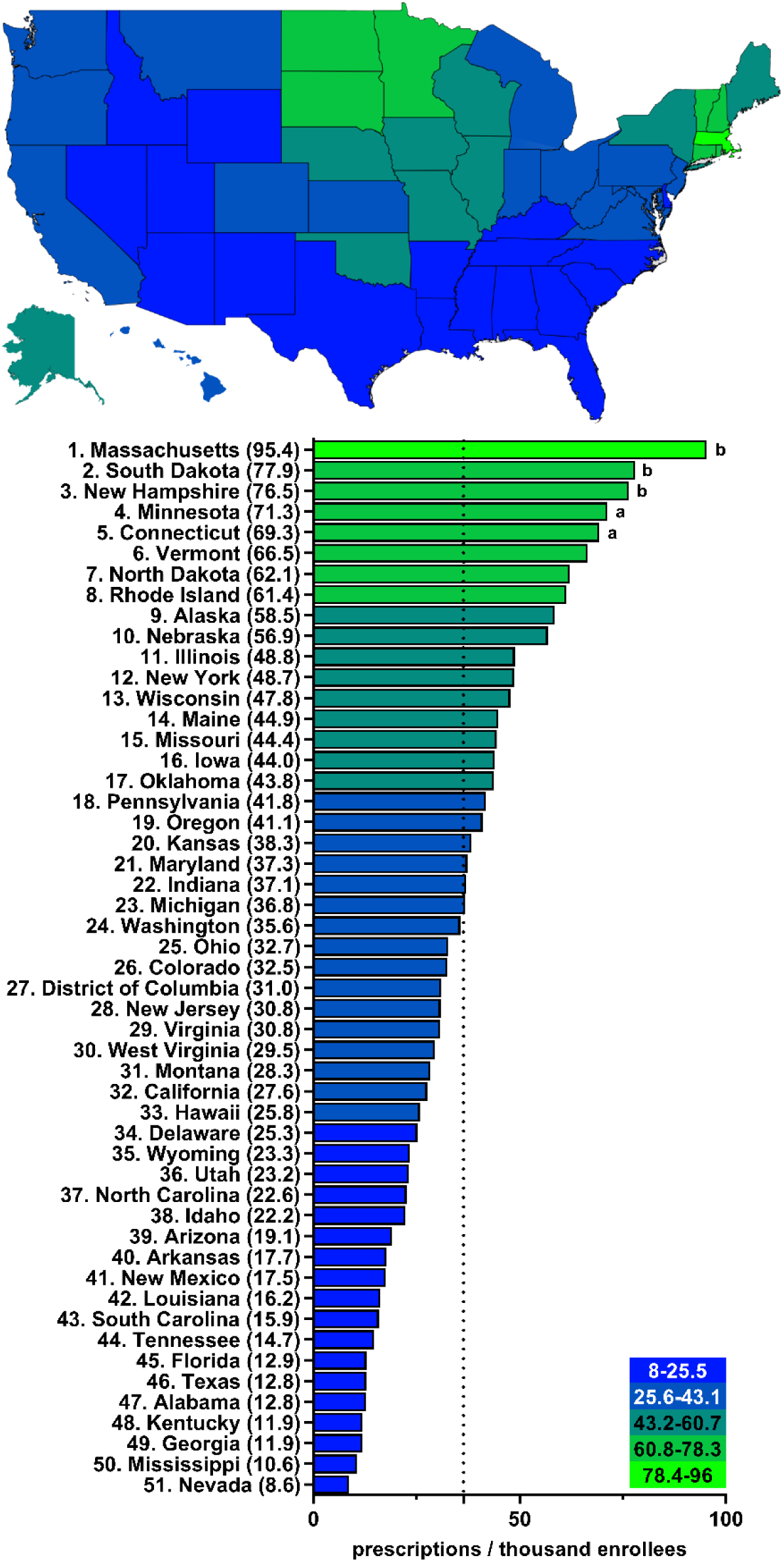
Large state-wide variation (11.1-fold) in number of clozapine prescriptions per thousand Medicare Part D enrollees in 2015 ^a^ indicates ≥1.50 SD (20.2) from the mean (36.4), which is denoted by the dotted line. ^b^ indicates ≥1.96 SD from the mean.

**Supplemental Figure 2.**
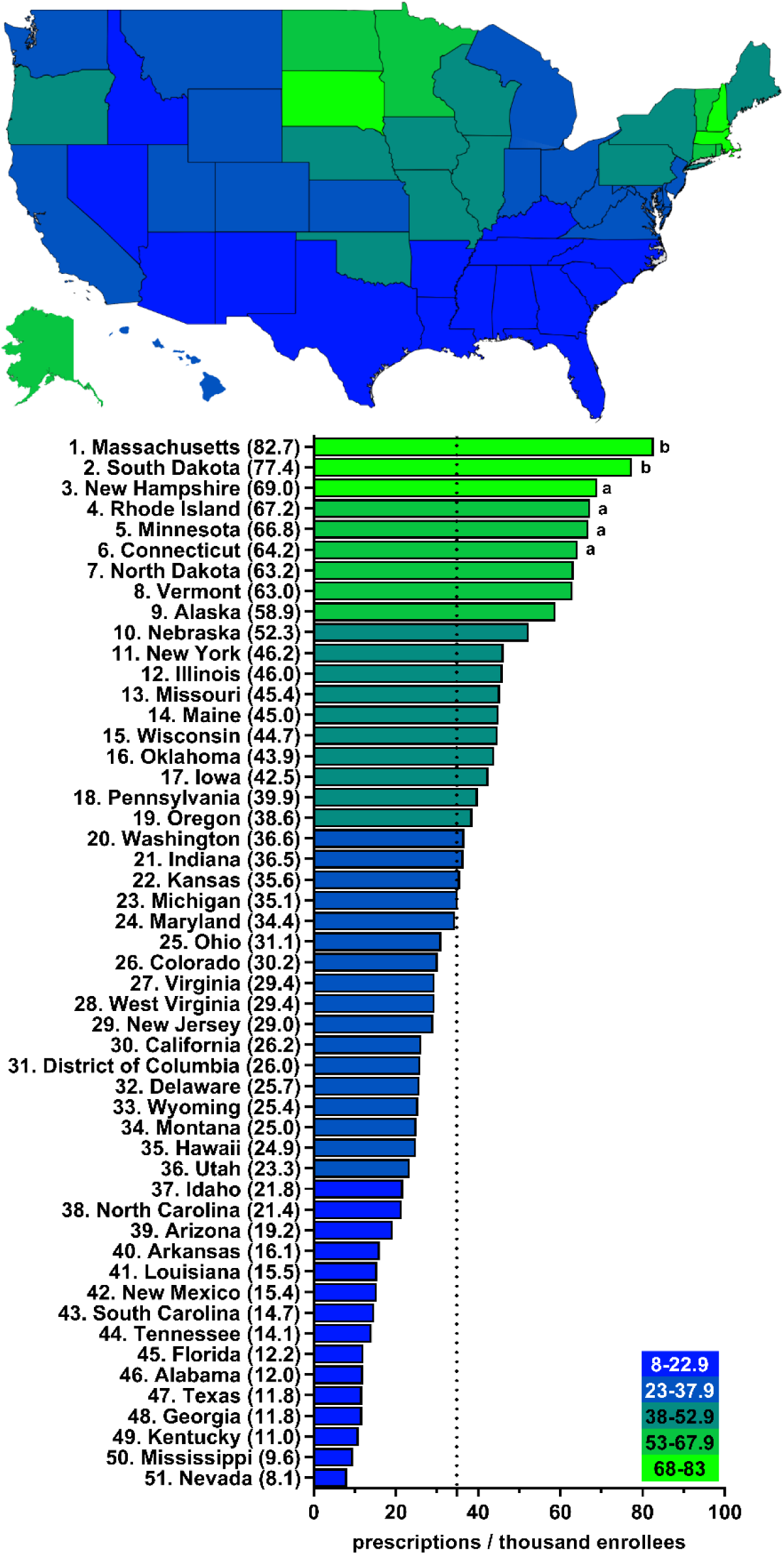
Large state-wide variation (10.2-fold) in number of clozapine prescriptions per thousand Medicare Part D enrollees in 2016 ^a^ indicates ≥1.50 SD (19.2) from the mean (34.8), which is denoted by the dotted line. ^b^ indicates ≥1.96 SD from the mean.

**Supplemental Figure 3.**
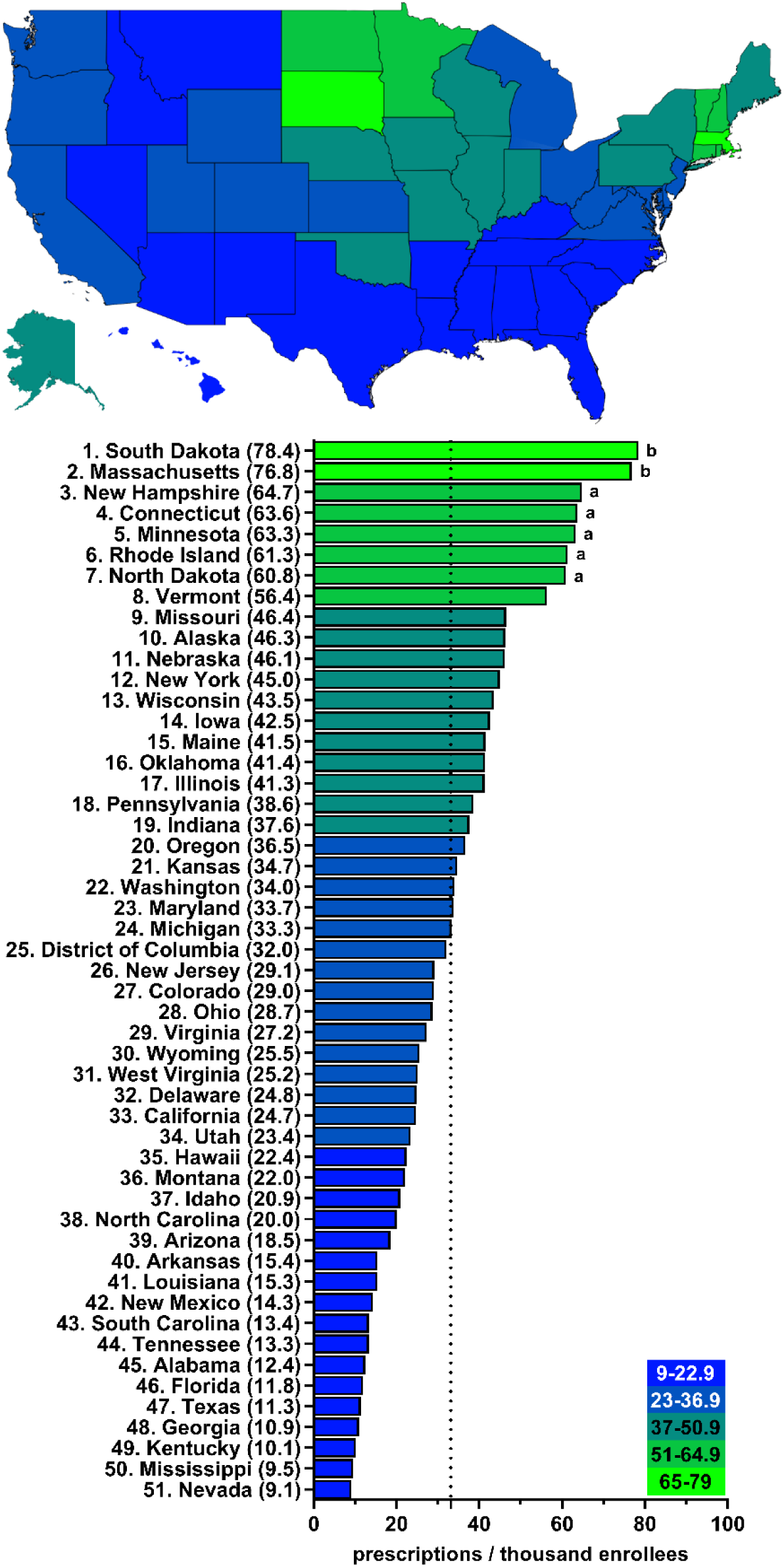
Large state-wide variation (8.6-fold) in number of clozapine prescriptions per thousand Medicare Part D enrollees in 2017 ^a^ indicates ≥1.50 SD (18.0) from the mean (33.1), which is denoted by the dotted line. ^b^ indicates ≥1.96 SD from the mean.

**Supplemental Figure 4.**
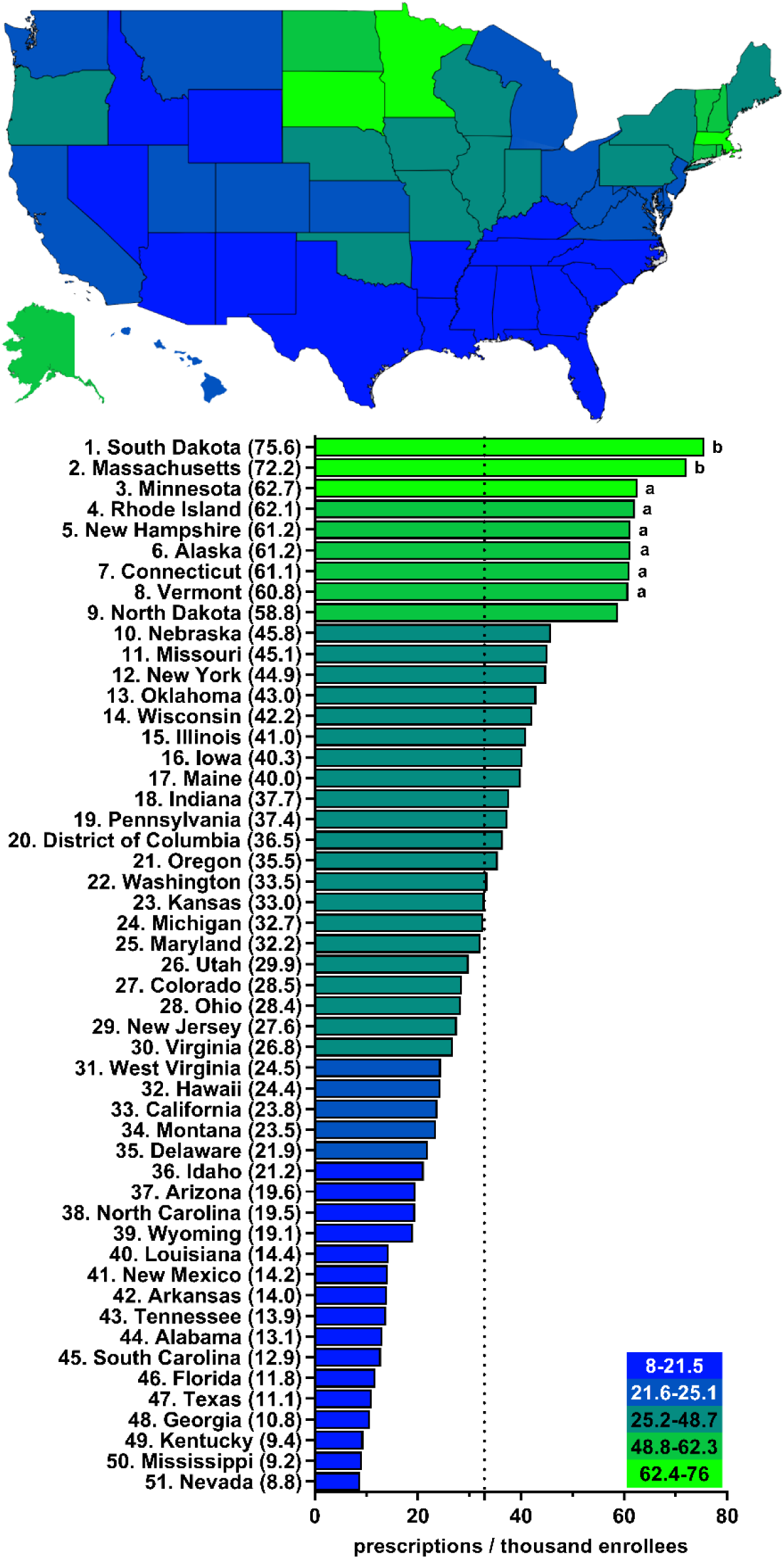
Large state-wide variation (8.6-fold) in number of clozapine prescriptions per thousand Medicare Part D enrollees in 2018 ^a^ indicates ≥1.50 SD (17.9) from the mean (32.9), which is denoted by the dotted line. ^b^ indicates ≥1.96 SD from the mean.

**Supplemental Figure 5.**
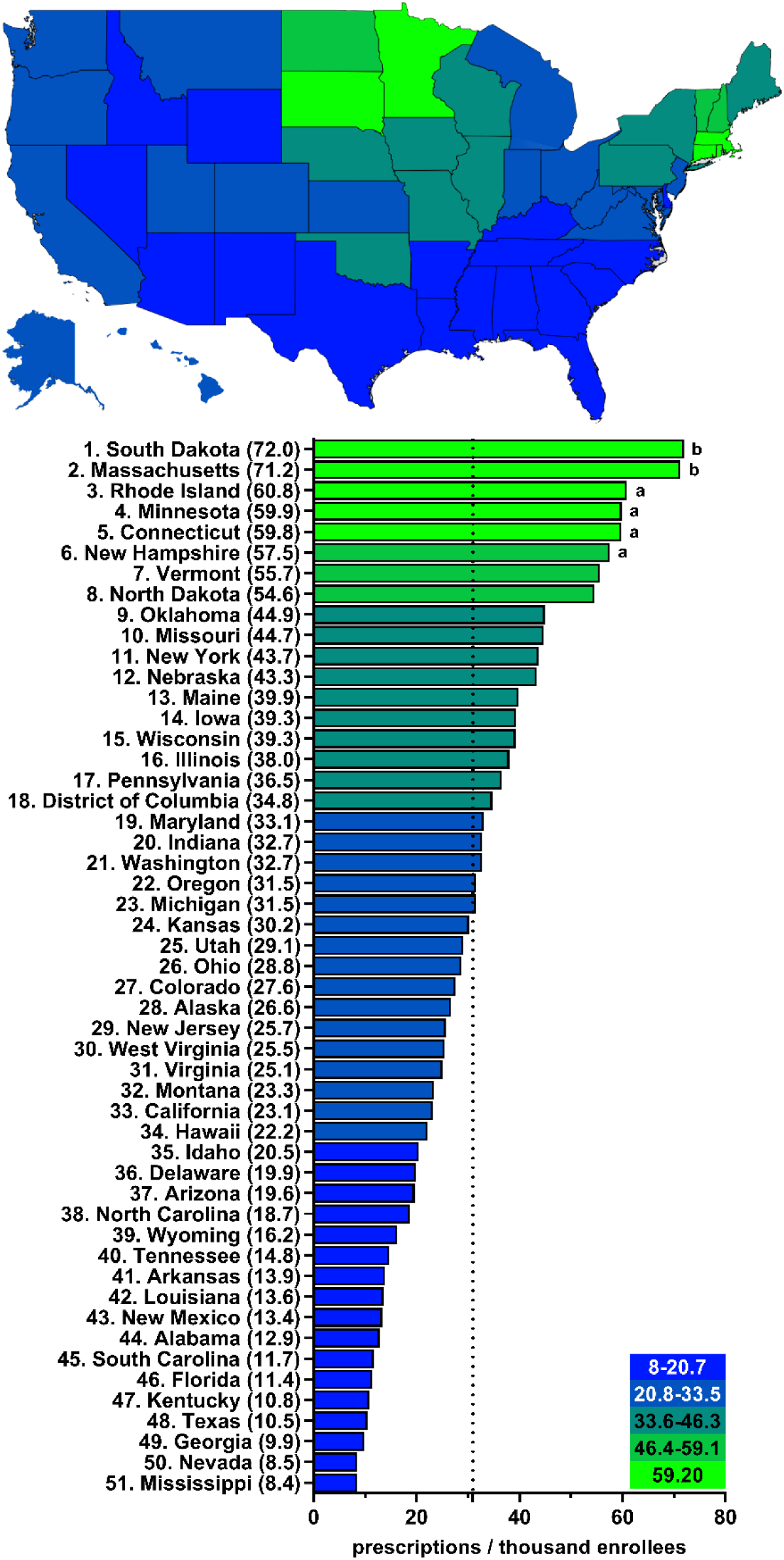
Large state-wide variation (8.5-fold) in number of clozapine prescriptions per thousand Medicare Part D enrollees in 2019 ^a^ indicates ≥1.50 SD (16.8) from the mean (31.0), which is denoted by the dotted line. ^b^ indicates ≥1.96 SD from the mean.

**Supplemental Figure 6.**
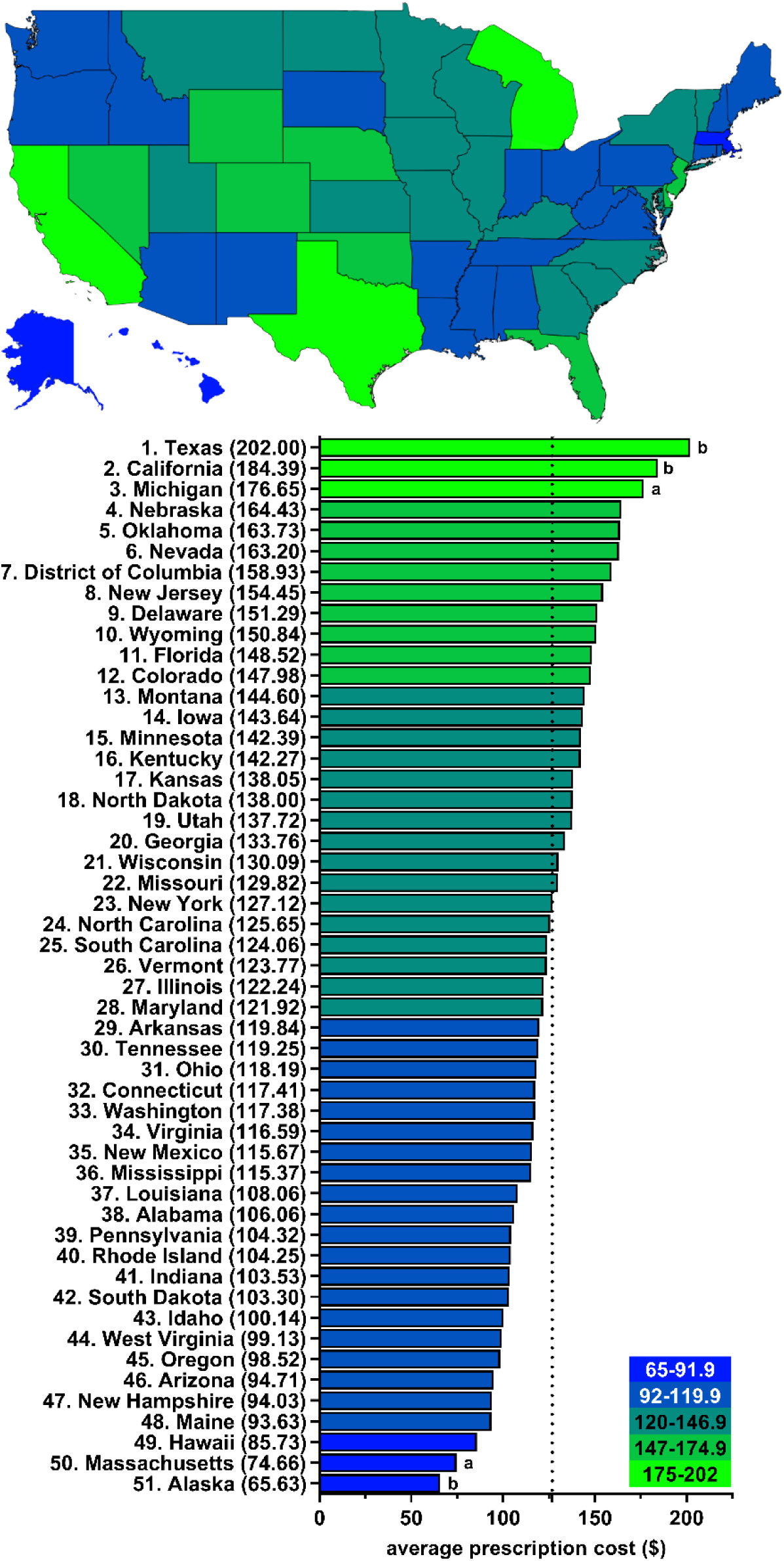
Pronounced state-wide variation (3.1-fold) in the average cost of a clozapine prescription for Medicare Part D enrollees in 2015. ^a^ indicates ≥1.50 SD ($27.39) from the mean ($126.80), which is denoted by the dotted line. ^b^ indicates ≥1.96 SD from the mean.

**Supplemental Figure 7.**
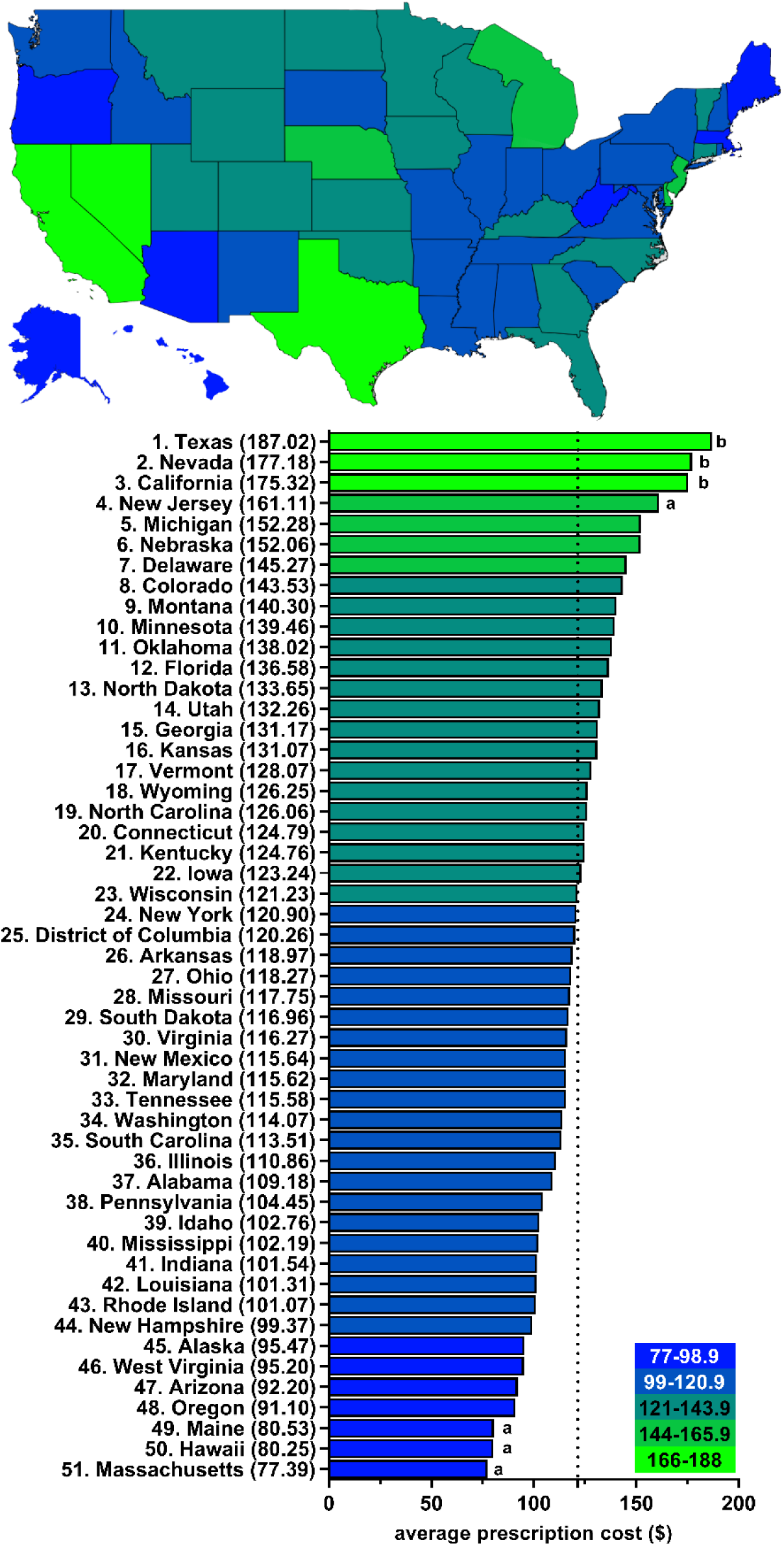
Pronounced state-wide variation (2.4-fold) in the average cost of a clozapine prescription for Medicare Part D enrollees in 2016. ^a^ indicates ≥1.50 SD ($23.56) from the mean ($121.56), which is denoted by the dotted line. ^b^ indicates ≥1.96 SD from the mean.

**Supplemental Figure 8.**
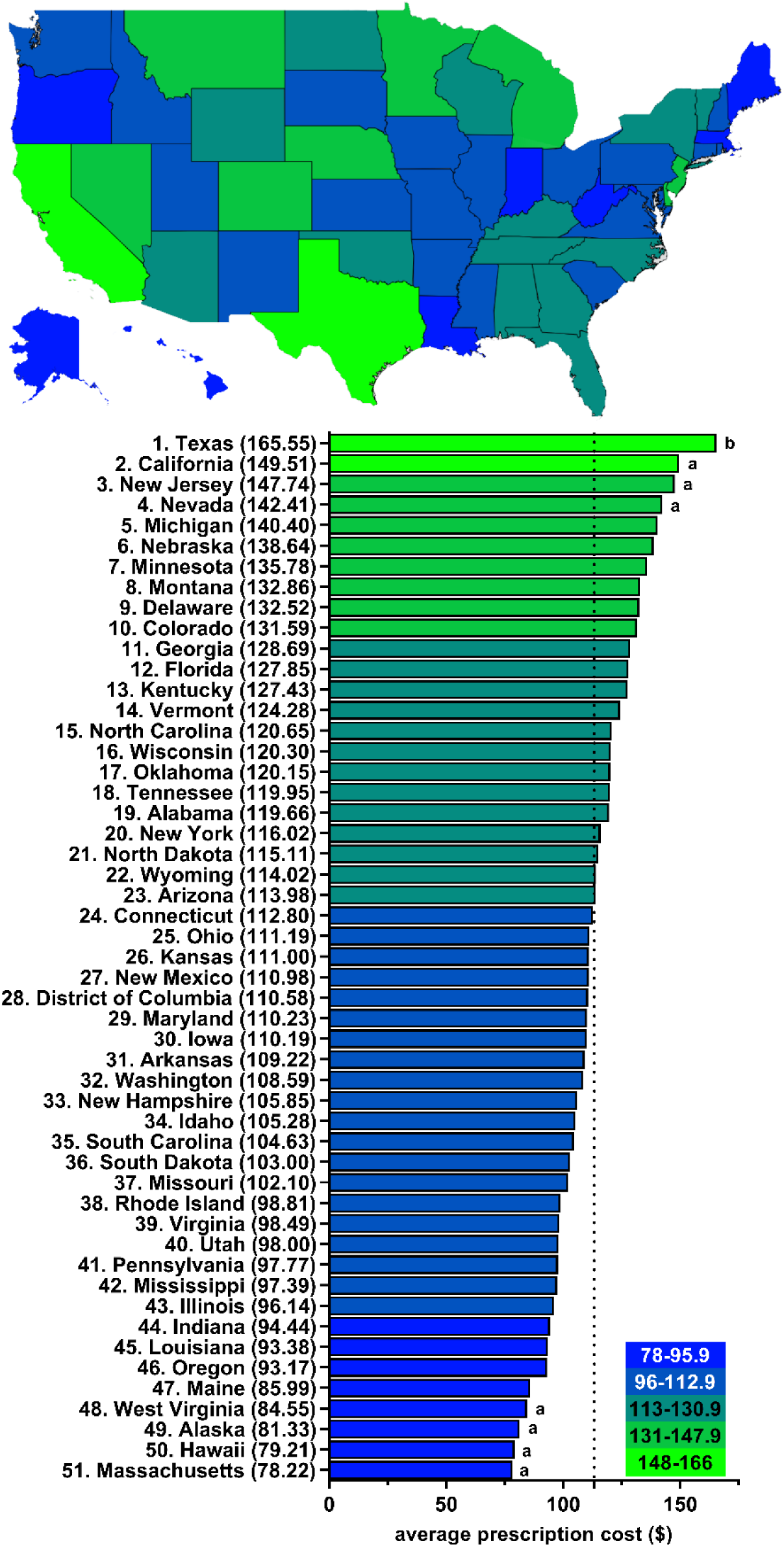
Pronounced state-wide variation (2.1-fold) in the average cost of a clozapine prescription for Medicare Part D enrollees in 2017. ^a^ indicates ≥1.50 SD ($18.87) from the mean ($113.29), which is denoted by the dotted line. ^b^ indicates ≥1.96 SD from the mean.

**Supplemental Figure 9.**
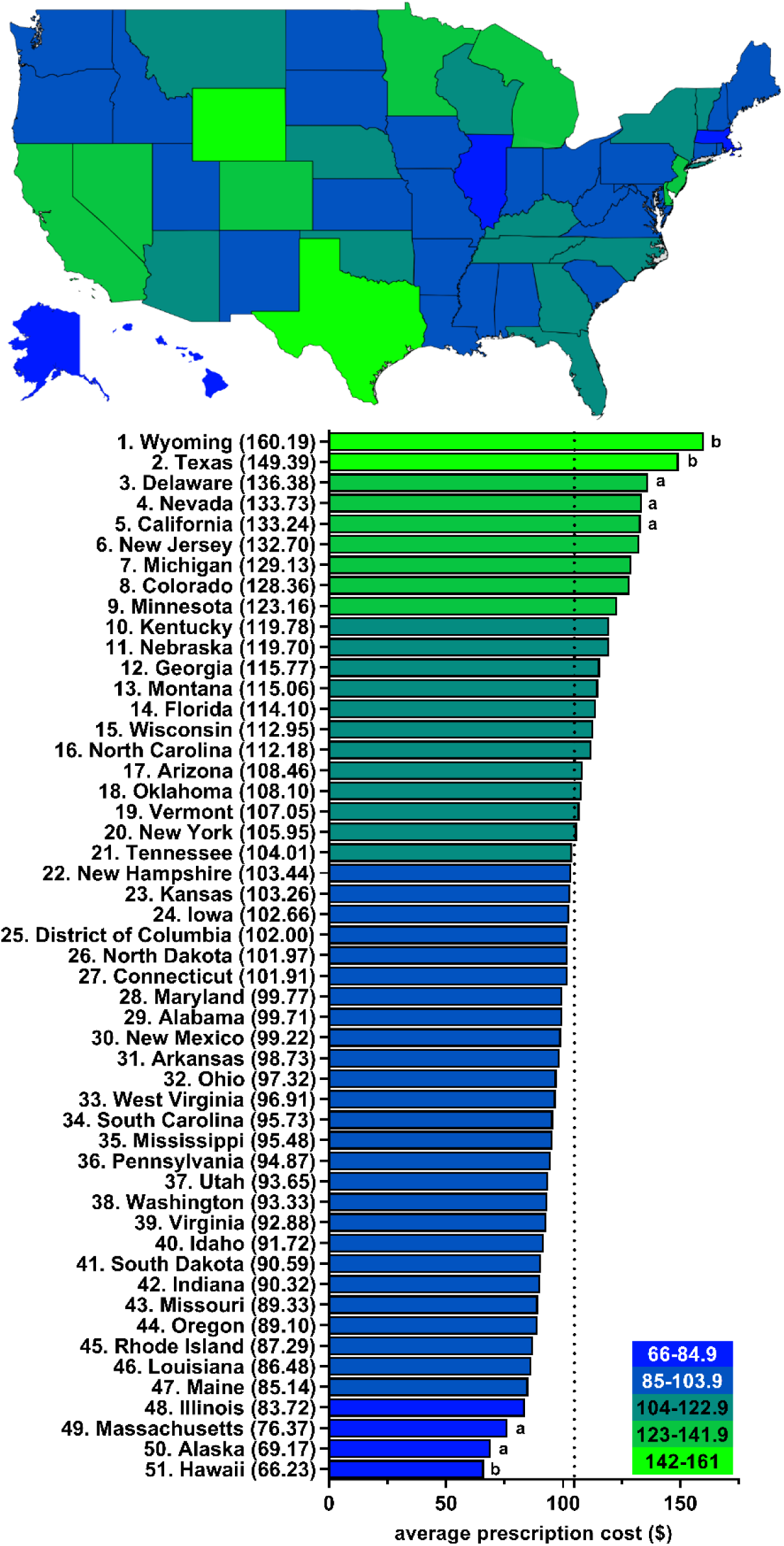
Pronounced state-wide variation (2.4-fold) in the average cost of a clozapine prescription for Medicare Part D enrollees in 2018. ^a^ indicates ≥1.50 SD ($18.74) from the mean ($104.86), which is denoted by the dotted line. ^b^ indicates ≥1.96 SD from the mean.

**Supplemental Figure 10.**
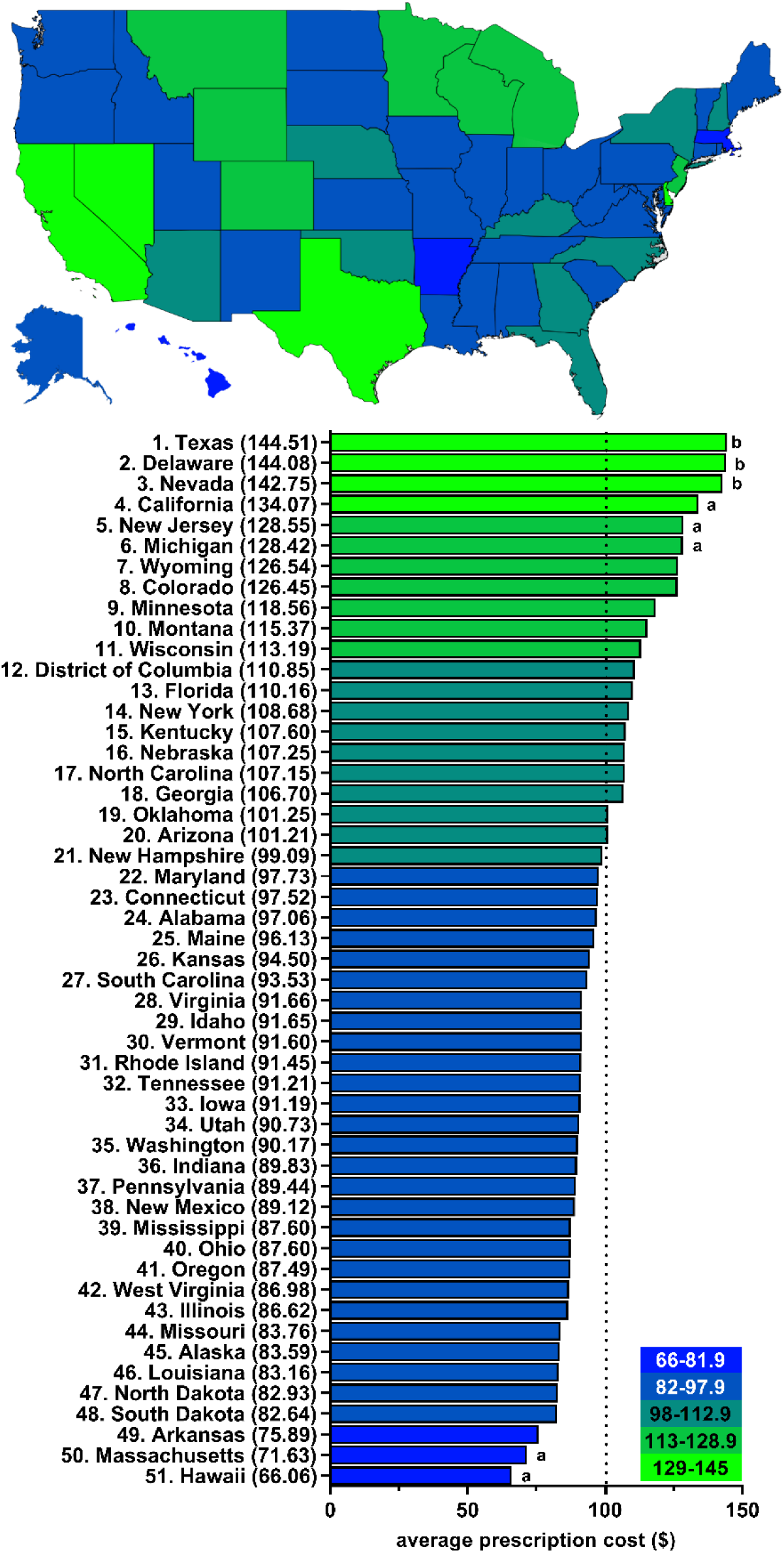
Pronounced state-wide variation (2.2-fold) in the average cost of a clozapine prescription for Medicare Part D enrollees in 2019. ^a^ indicates ≥1.50 SD ($18.26) from the mean ($100.45), which is denoted by the dotted line. ^b^ indicates ≥1.96 SD from the mean.

